# Cross-sectional analysis: interpretation of non-statistically significant results in randomised controlled clinical trials in rehabilitation

**DOI:** 10.1101/2024.09.16.24313294

**Authors:** Caterina Mugnai, Luca Falsiroli Maistrello, Giacomo Fiacca, Michele Perucchini, Noemi Corbetta, Federico Amateis, Stefano Salvioli

## Abstract

**Introduction:** Despite the CONSORT guidelines, which aim to improve the quality of studies, authors often formulate conclusions based on the dichotomous distinction of the p-value, declaring differences between ‘statistically significant’ and ‘non-significant’. This approach confuses the identification of the real efficacy of the studied treatment. To solve this problem, CONSORT guidelines recommend using confidence intervals, which offer a more complete view of possible effects. However, authors’ conclusions often remain based on a binary approach, confusing the absence of evidence with the evidence of absence. This error can influence clinical practice and future research, leading to the identification of ‘negative’ treatments based on ‘statistical insignificance’, which reflects a lack of evidence of absence, not the absence of evidence.

**Objectives:** To assess the prevalence of misinterpretation of non-statistically significant results, both in the abstract and in the article, in a sample of all randomised controlled trials (RCTs) with non-statistically significant primary outcomes published in 5 rehabilitation journals with the highest impact factor (IF) published between 2019 and 2023 and to assess whether the primary outcome result is reported according to CONSORT guidelines.

**Methods:** We will conduct a cross-sectional analysis of all Rcts with non-statistically significant primary outcomes in 5 general rehabilitation journals with the highest IF published between 2019 and 2023. We will determine the prevalence of trials in which non-significance is interpreted as absence of evidence, evidence of absence, or advice to use the intervention in clinical practice in the abstract and article conclusions, and the prevalence of trials that adhered to CONSORT guidelines for reporting the primary outcome.

## INTRODUCTION

The scientific literature and the Evidence-Based Medicine (EBM) approach emphasise the necessity of developing scientific evidence based on rigorously conducted trials. The randomised controlled trial (RCT) is regarded as the most reliable type of study for guiding and legitimising treatment decisions^1^. The importance of RCTs is also supported by the GRADE approach which, in systematic reviews with meta-analysis, classifies the evidence from this study design with a high quality of evidence ^2^.

To improve reporting quality, CONsolidated Standards Of Reporting Trials (CONSORT) guidelines^2^ have been developed. The need to pre-define the primary and secondary outcomes of the trial to minimise the possibility of serendipitous discoveries is one of the key messages of these guidelines^3^. As indicated by the American Statistical Association in 2016, a rigorous process of data analysis and interpretation of the results is of central importance in reaching conclusions that can be considered reliable^4^.

Nevertheless, authors frequently formulate statements based on a dichotomous distinction of the p-value index. Differences in outcome results between the two study groups are declared “statistically significant” when the p-value is below a cut-off set a priori (generally 0.05) and “not statistically significant” when equal or greater. This approach confuses an element relating to the usual/unusual presentation of the data linked to the statistical test with a characteristic of the effect and the population analysed^5,6^.

In order to address this misinterpretation, the CONSORT guidelines emphasise the importance of reporting confidence intervals, which shift the focus from a single point value to the entire range of effect sizes.^6^ Despite the significance of interpreting confidence intervals and the information they can provide for both clinical practice and future research, author conclusions are often linked to a binary approach related to the distribution of the data with respect to the null value, which falls under the same concept of ‘statistical significance’. In particular, the issue arises when inconclusive studies define a treatment as ineffective. This is known as confounding the absence of evidence of a difference with evidence of no difference^7^.

The consequences of such interpretative errors are twofold. From the clinical perspective, they have the potential to impact future scientific research and the interventions chosen for patients. From the statistical perspective, they may result in the identification of ‘ negative ’ treatments within RCTs based on ‘statistical insignificance’^8^. This definition is inaccurate in suggesting that the trial concluded that there was no difference, when in fact it indicates a lack of evidence of a difference.^7^

It is possible to observe a phenomenon within studies that have produced non-statistically significant results, which is known as ‘spin’. This is characterised by the reporting of data on primary outcomes that are not statistically significant. It is achieved through the use of misleading linguistic strategies that distort the conclusions that should be drawn from the data.^8^ These reporting modalities, whether consciously or unconsciously employed, may include the deliberate omission of data, the accentuation of the efficacy of a treatment or the comparability of two treatments despite non-statistical significance, the distraction from the lack of significance of the primary outcome, the concentration on secondary outcomes or the emphasis on intra-group improvements instead of inter-group comparison^9,10^. Number of studies have been conducted in the literature with the objective of analysing the presence of spin in articles pertaining to various clinical fields, including urology^11^,bariatric surgery^12^, oncology^13^, plastic surgery^14^ and dental caries^15^

In the context of rehabilitation, an illustrative example is the RCT by Akbari et al., published in 2016. Despite the lack of statistical significance, the authors recommend the use of balance training in subjects who have undergone ACL reconstruction. However, the conclusions and language used in the study are inaccurate and misleading, which detracts from the non-significance of the results: “[…] balance exercise could partially improved dynamic stability index in early stage of ACL reconstruction rehabilitation.”^16^

A 2022 meta-analysis by Hemming et al.revealed that even in high-impact factor general medical journals, articles with erroneous conclusions regarding the results obtained are published.^16^

It is therefore essential that readers are made aware of the importance of correctly interpreting scientific studies by analysing them with a more critical point of view.

For these reasons, the purpose of this review is to assess:

1. the prevalence of misinterpretation of non-statistically significant results, both in the abstract and in the article, in all RCTs with non-statistically significant primary outcomes published in 5 rehabilitation journals with the highest IF in the rehabilitation section, published between 2019 and 2023. In particular, if:
  - non-significance is interpreted as evidence of absence (i.e. evidence of no difference);
  - non-significance is nevertheless interpreted as advice to use that intervention;
  - the width of the confidence interval is interpreted considering a possible Beta error;
2. whether the primary outcome result is reported in accordance with the CONSORT guideline, i.e. in the form of a point estimate (mean difference for continuous outcomes and both absolute and relative risks for dichotomous outcomes) and its confidence interval.

## OBJECTIVES

Our primary objective will be to assess the prevalence of misinterpretation of non-statistically significant results, both in the abstract and in the article, in all RCTs with non-statistically significant primary outcomes published in 5 rehabilitation journals with the highest IF in the rehabilitation section, published between 2019 and 2023.

In particular if:

- non-significance is interpreted as evidence of absence (i.e. evidence of no difference);
- non-significance is nevertheless interpreted as advice to use that intervention;
- the width of the confidence interval is interpreted considering a possible Beta error.

As a secondary outcome, we will assess whether the primary outcome result is reported in accordance with the CONSORT guideline ^17^, i.e. in the form of a point estimate (mean difference for continuous outcomes and both absolute and relative risks for dichotomous outcomes) and its confidence interval.

## METHODS AND ANALYSIS

We will conduct a cross-sectional analysis of all RCTs with non-statistically significant primary outcomes published between 2019 and 2023 in 5 generic rehabilitation journals with the highest IF according to the InCites Journal Citation Report of 2022^18^. The choice of years and number of journals was based on the study by Hemming et al^19^.

Below are the five selected journals indexed in InCites:

- *Journal of Physiotherapy (2022 IF 12,1)*
- *Journal of Orthopaedic & Sports Physical Therapy (2022 IF: 6,1)*
- *Journal of NeuroEngineering and Rehabilitation (2022 IF: 5,1)*
- *IEEE Transactions of Neural Systems and Rehabilitation Engineering (2022 IF: 4,8)*
- *Annals of Physical and Rehabilitation Medicine (2022 IF: 4,6)*

### Eligibility criteria

Inclusion criteria:

- All RCTs with non-statistically significant primary outcomes published between 2019 and 2023 in 5 generic rehabilitation journals with the highest IF according to the InCites Journal Citation Report of 2022^18^ will be included.

Exclusion criteria:

- RCTs with statistically significant primary outcomes, protocols or secondary analysis articles will be excluded.
- RCTs with multiple primary outcomes and at least one of them statistically significant will be excluded.
- RCTs with multiple outcomes without defining the primary will be excluded.

### Study selection process

We have identified the journal tags for the 5 journals in Medline (PubMed) and a detailed search strategy to find all RCTs published from 2019 to 2023 by using the INTER-Tasc search filter resource ^20^.

> *((((((J Physiother) OR (J Orthop Sports Phys Ther)) OR (J Neuroeng Rehabil)) OR (ieee trans neural syst rehabil eng)) OR (ann phys rehabil med))) AND ((“randomized controlled trial”[PT] OR “controlled clinical trial”[Publication Type] OR “randomized”[Title/Abstract] OR “placebo”[Title/Abstract] OR “drug therapy”[MeSH Subheading] OR (“randomly”[Title/Abstract] OR “trial”[Title/Abstract] OR “groups”[Title/Abstract])) NOT (“animals”[MeSH Terms] NOT “humans”[MeSH Terms])) Filters: from 2019/1/1 - 2023/12/31*

The search will be only conducted on Medline because it indexes all considered journals. The identified records will undergo a two-stage selection process, the first by reading the title and abstract using Rayyan online software ^21^, and the second by reading the full text. Both stages will be carried out by two authors (MP and GF) independently. Each of these will select potentially suitable articles based on predetermined criteria and any discrepancies will be resolved with the help of a third reviewer (CM). The study selection process will be summarised in a flow chart.

### Data extraction

A data extraction form will be used, and the following data will be extracted:

- first author and year of publication;
- whether the studies provided adequate justification for their conclusions in the abstract:
  - non-significance is interpreted as absence of evidence (i.e. the correct interpretation for a negative result); and, in this case, if the width of the confidence interval is interpreted, for example by considering a possible Beta error (i.e. recognizing some uncertainty due to low sample size).
  - non-significance is interpreted as evidence of absence (i.e. evidence of no difference);
  - non-significance is still interpreted as advice to use that intervention in clinical practice;
- whether the studies provided adequate justification for their conclusions in the article:
  - non-significance is interpreted as absence of evidence (i.e. the correct interpretation for a negative result); and, in this case, if the width of the confidence interval is interpreted, for example by considering a possible Beta error (i.e. recognizing some uncertainty due to low sample size).
  - non-significance is interpreted as evidence of absence (i.e. evidence of no difference);
  - non-significance is still interpreted as advice to use that intervention in clinical practice;
- adherence to CONSORT guidance on reporting of the primary outcome (i.e., in the form of point estimate [mean difference for continuous outcomes and absolute and relative risks for dichotomous outcomes] and associated confidence interval) or not;

Data extraction will be performed by two reviewers (FA and NC) independently and, when necessary, a third author (CM) will solve disagreements.

### Data analysis

Count data and percentage of studies for each of the following outcomes will be provided:

1. prevalence of studies where the non-significance is interpreted as an absence of evidence (i.e. the correct interpretation for a negative result);
  a. and, in this case, if the width of the confidence interval is interpreted, for example by considering a possible Beta error (i.e. recognizing some uncertainty due to low sample size).
2. prevalence of studies where the non-significance is misinterpreted in the abstract;
  a. prevalence of studies where the non-significance is interpreted as evidence of absence (i.e. evidence of no difference) in the abstract;
  b. prevalence of studies where the non-significance is still interpreted as advice to use that intervention in clinical practice in the abstract;
3. prevalence of studies where the non-significance is misinterpreted in the article:
  a. prevalence of studies where the non-significance is interpreted as evidence of absence (i.e. evidence of no difference) in the article;
  b. prevalence of studies where the non-significance is still interpreted as advice to use that intervention in clinical practice in the article;
4. prevalence of studies where the adherence to CONSORT guidance on reporting of the primary outcome (i.e., in the form of point estimate [mean difference for continuous outcomes and absolute and relative risks for dichotomous outcomes] and associated confidence interval) is satisfied.

## Data Availability

All data produced in the present work are contained in the manuscript

## ETHICS AND DISSEMINATION

An article with the results obtained will be prepared and submitted for publication in a peer-reviewed journal of the rehabilitation field. The results will also be disseminated at a relevant international conference.

## AUTHORS’ CONTRIBUTIONS

The study protocol was conceived and designed by CM, LFM and SS. GF, MP, CM and SS developed the search strategy. CM, LFM, SS, GF, MP, FA, and NC contributed to conceptualizing the study objectives, refining the search strategy, establishing study selection criteria, and outlining plans for data extraction. The final version of the protocol was approved by CM, LFM, and SS.

## FUNDING STATEMENT

This research received no specific grant from any funding agency in the public, commercial or not-for-profit sectors.

## Notes

### Competing Interest Statement

The authors have declared no competing interest.

### Funding Statement

This study did not receive any funding

